# Identifying body awareness-related brain network changes after Spring Forest Qigong™ practice or P.Volve low-intensity exercise in adults with chronic low back pain: a feasibility Phase I Randomized Clinical Trial

**DOI:** 10.1101/2023.02.11.23285808

**Authors:** Ann Van de Winckel, Lin Zhang, Timothy Hendrickson, Kelvin O. Lim, Bryon A. Mueller, Angela Philippus, Kimberley R. Monden, Jinseok Oh, Qiyin Huang, Jacquelyn V.L. Sertic, Jacquelyn Ruen, Jürgen Konczak, Roni Evans, Gert Bronfort

## Abstract

**Background:** Chronic low back pain (cLBP) affects the quality of life of 52 million Americans and leads to an enormous personal and economic burden. A multidisciplinary approach to cLBP management is recommended. Since medication has limited efficacy and there are mounting concerns about opioid addiction, the American College of Physicians and American Pain Society recommend non-pharmacological interventions, such as mind and body approaches (e.g., Qigong, yoga, Tai Chi) before prescribing medications. Of those, Qigong practice might be most accessible given its gentle movements and because it can be performed standing, sitting, or lying down. The three available Qigong studies in adults with cLBP showed that Qigong reduced pain more than waitlist and equally well than exercise. Yet, the duration and/or frequency of Qigong practice were low (<12 weeks or less than 3x/week). The objectives of this study were to investigate the feasibility of practicing Spring Forest Qigong™ or performing P.Volve low intensity exercises 3x/week for 12 weeks, feasibility of recruitment, data collection, delivery of the intervention as intended, as well as identify estimates of efficacy on brain function and behavioral outcomes after Qigong practice or exercise. To our knowledge, this is the first study investigating the feasibility of the potential effect of Qigong on brain function in adults with cLBP.

**Methods:** We conducted a feasibility Phase I Randomized Clinical Trial. Of the 36 adults with cLBP recruited between January 2020 and June 2021, 32 were enrolled and randomized to either 12 weeks of remote Spring Forest Qigong™ practice or remote P.Volve low-intensity exercises. Participants practiced at least 3x/week for 41min/session with online videos. Our main outcome measures were the Numeric Pain Rating Scale (highest, average, and lowest cLBP pain intensity levels in the prior week), assessed weekly and fMRI data (resting-state and task-based fMRI tasks: pain imagery, kinesthetic imagery of a Qigong movement, and robot-guided shape discrimination). We compared baseline resting-state connectivity and brain activation during fMRI tasks in adults with cLBP with data from a healthy control group (n=28) acquired in a prior study. Secondary outcomes included measures of function, disability, body awareness, kinesiophobia, balance, self-efficacy, core muscle strength, and ankle proprioceptive acuity with a custom-build device.

**Results:** Feasibility of the study design and methods was demonstrated with 30 participants completing the study (94% retention) and reporting high satisfaction with the programs; 96% adherence to P.Volve low-intensity exercises, and 128% of the required practice intensity for Spring Forest Qigong™ practice. Both groups saw promising reductions in low back pain (effect sizes Cohen’s *d*=1.01-2.22) and in most other outcomes (*d*=0.90-2.33). Markers of ankle proprioception were not significantly elevated in the cLBP group after the interventions. Brain imaging analysis showed weaker parietal operculum and insula network connectivity in adults with cLBP (n=26), compared to data from a healthy control group (n=28). The pain imagery task elicited lower brain activation of insula, parietal operculum, angular gyrus and supramarginal gyrus at baseline in adults with cLBP than in healthy adults. Adults with cLBP had lower precentral gyrus activation than healthy adults for the Qigong movement and robot task at baseline. Pre-post brain function changes showed individual variability: Six (out of 13) participants in the Qigong group showed increased activation in the parietal operculum, angular gyrus, supramarginal gyrus, and precentral gyrus during the Qigong fMRI task.

**Interpretation:** Our data indicate the feasibility and acceptability of using Spring Forest Qigong™ practice or P.Volve low-intensity exercises for cLBP relief showing promising results in terms of pain relief and associated symptoms. Our brain imaging results indicated brain function improvements after 12 weeks of Qigong practice in some participants, pointing to the need for further investigation in larger studies.

**Trial registration number:** ClinicalTrials.gov: NCT04164225.

## 1. INTRODUCTION

Low back pain is a leading cause of disability worldwide.^1–3^ The lifetime prevalence of low back pain is about 40% with 5-10% of acute pain developing into chronic low back pain (cLBP), defined as having low back pain for longer than 3 months.^4^ About 85% of adults with cLBP have non-specific low back pain, where an underlying pathology or a nociceptive contributor cannot yet been identified.^5^ cLBP is thus defined based on symptoms, and includes pain, muscle tension, or stiffness localized below the costal margin and above the inferior gluteal folds, with or without sciatica (i.e., pain traveling down the leg from the lower back).

cLBP has become widely recognized as a biopsychosocial condition^6,7^ and is often associated with functional disability, more work absences, more severe feelings of depression and anxiety, and reduced quality of life.^4,8^ Decreased lumbar and core muscle strength, balance, endurance, altered body awareness/mental body representations, and lumbar proprioception can increase pain intensity and disability in adults with non-specific cLBP.^9–16^ Additionally, chronic pain can be perpetuated by decreased emotional coping and increased avoidance of moving for fear of pain. ^10,17^

Low back pain is the most common reason for seeking care in the healthcare system.^18–20^ The utilization of health care services for chronic LBP has increased substantially over the past two decades, with opioid use as one of the treatments, despite the associated risks and limited evidence of improved function or reduced pain.^18–20^ In 2016, low back and neck pain had the highest amount of health care spending with an estimated $134.5 billion in spending.^21^ The CDC Clinical Practice Guideline for Prescribing Opioids for Pain — United States, 2022^19,22^ as well as the American College of Physicians and American Pain Society ^23^ recommend that non-pharmacological interventions, such as physical therapy, exercise, and mind and body approaches (e.g., Qigong, yoga, Tai Chi) be selected prior to prescribing medications (non-steroidal anti-inflammatory drugs or duloxetine).^24^

There is moderate evidence that exercise is better than no treatment, medication, or placebo comparisons.^25^ A recent systematic review identified the most common type of exercises investigated for cLBP: core strengthening (30%), programs with three or more types of exercises (26%), and general strengthening exercises (12%). About 45% of exercise treatments were ‘back specific’ and 29% were ‘whole body’ exercises.^26^ Hayden *et al*. (2021) found that Pilates, McKenzie therapy, and functional restoration were more effective than other types of exercise treatment for reducing pain intensity and functional limitations.^26^

Mind and body approaches are suited to address both the mind and the body aspects, and thus could be good candidates to address the biopsychosocial aspects of chronic pain.^27^ Of those, Qigong practice might be most accessible given its gentle movements and because it can be performed standing, sitting, or lying down. So far, Qigong has been investigated in three studies and showed that Qigong reduced pain more than waitlist^28,29^ and equally well than exercise.^30^ Yet, the duration and/or frequency of Qigong practice were low (<12 weeks or less than 3x/week).^28–30^

The benefits of Qigong over exercise might be the combined present-moment body awareness with gentle movements versus a primary focus on physical movements during exercise, a difference that may be identified and quantified with brain imaging. Previous research in mindfulness and focused breathing meditation showed that the insula regulates the autonomic nervous system, nociception, and interoception, such as during attentive breathing.^31–38^ Insula provides a meta-representation of the entire body state by integrating internal body sensations (i.e., interoception) and sensations perceived with e.g. touch or vision (i.e., exteroception).^39^ The insula receives input from the parietal operculum. The parietal operculum is part of the multimodal integration network that links body awareness to visuospatial body maps (in the posterior parietal cortex) to guide motor actions.^40–44^ Body sensations processed in parietal operculum are sent to the insula and its connections (i.e., interoceptive network), and then from the insula to the prefrontal cortex, bringing body awareness into conscious awareness.^31,39^ Additionally, the parietal operculum and insula are also known to be key areas for pain perception.^45^ Based on these data, we believe that Qigong similarly activates the parietal operculum and the insula and its interoceptive network.

Therefore, the objectives of this study were to investigate the feasibility of practicing Spring Forest Qigong™ or performing P.Volve low intensity exercises 3x/week for 12 weeks, feasibility of recruitment, data collection, delivery of the intervention as intended, as well as identifying estimates of efficacy on brain function and behavioral outcomes after Qigong practice or exercise.^46,47^ There is evidence in adults with fibromyalgia that exercise of moderate intensity can influence descending regulatory systems,^48^ but to the best of our knowledge, effects of either Qigong or exercise on brain function in adults with cLBP have not yet been investigated.

## 2. METHODS

### 2.1 Study design

We performed a feasibility Phase I Randomized Clinical Trial in community-dwelling adults with cLBP in Minnesota, US. Written informed consent was obtained. The study was conducted in accordance with the principles of the Declaration of Helsinki (2013)^49^ and the Institutional Review Board (IRB) of the University of Minnesota approved the study (STUDY00005656). The CONSORT reporting guidelines were followed.^50,51^ Given that this was a Phase I randomized clinical trial, there was no formal data monitoring committee, but a data integrity monitor of the University of Minnesota performed monitoring every 6 months for the duration of the study.

### 2.2 Participants

We included adults with cLBP (target n=32, based on sample size calculation detailed below), without restriction for race, sex or socio-economic status. Participants were recruited using multiple approaches, including postings on StudyFinder, a website managed by the UMN CTSI’s Recruitment Center; identification via UMNs Research Match, an electronic volunteer recruitment registry; postings in the University of Minnesota clinics, local rehabilitation centers, on relevant websites and on professional social media sites. Additionally, we capitalized on ongoing recruitment efforts in a NIH funded large (n=1200), multi-year study of acute and subacute LBP (<u>UH3AT008767-02</u>) led by one of our co-authors (Bronfort), where large numbers of patients with LBP were identified during the screening process. Adults with cLBP who were excluded from that study were invited to consider participation in the present study.

Inclusion criteria were adults aged 18-75 years, with non-specific LBP (i.e., without radiating pain or neurological signs) lasting >3 months, average LBP severity ≥ 4 on the 0-10 numerical pain rating scale (NPRS) over the past week; willing to participate in a Qigong or exercise intervention for 12 weeks; read and write fluently in English.

Exclusion criteria included (1) specific causes of LBP such as spinal fracture, infection, cancer, spinal stenosis, disc herniation, cauda equina syndrome; (2) progressive neurological deficits, inflammatory spinal arthropathy, surgical fusion of the lumbar spine; (3) active management of current LBP episode by another healthcare provider; (4) regular practice of Tai Chi, Qigong, yoga, or exercise in the past year; (5) adults with MRI-contra-indications (6) pregnant; (7) untreated serious mental health disorders (e.g. major depressive or psychotic disorders); (8) substance abuse; (9) decreased cognitive function (Mini-Mental State Examination-short version <13/16); (10) inability or unwillingness to give written informed consent.

Demographic data including age, sex (female, male, other), gender identification (free text), race (American Indian or Alaska Native, Asian, Black or African-American, Native Hawaiian or other Pacific Islander, White, unknown, not reported), and ethnicity (Hispanic, non-Hispanic, unknown, not reported) were collected through self-report.

For the baseline comparison of brain imaging of adults with cLBP with healthy adults, we used brain imaging data from 28 healthy adults from a previous study.^52^

### 2.3 Screening

After signing the HIPAA and consent form, baseline characteristics were collected: demographics, general health data (including medical history), past healthcare use, scoring of low back pain intensity in the week prior to the first clinical assessment, the National Institutes of Health Task Force on Research Standards for cLBP with minimum recommended NINDS-CDE dataset and the Quebec Task Force classification of spinal disorders.

To ensure safety, an in-person physical screening by experienced physical therapists with years of practice was performed in the first author (VDW)’s Brain Body Mind Lab, at the University of Minnesota. The clinical screening entailed the Romberg test to detect balance dysfunction, the Straight Leg Raise test to 45 degrees to detect signs of nerve root tension, and a neurological exam (reflexes, sensation, muscle strength). The Mini-Mental State Examination-Brief version (exclusion for <13/16) was used to assess cognitive impairments.

### 2.4 Randomization and masking

We used computer-generated randomization. There was no stratification. Participants were randomized into two groups: GROUP A was practicing Spring Forest Qigong™ “5 Element Healing Movements” for 12 weeks; GROUP B was practicing P.Volve low-intensity exercises for 12 weeks. Both groups received an in-person or online introduction class (online introduction classes were inserted during the acute period of the COVID-19 pandemic) and then practiced at home with an online video. The frequency and duration of 12 weeks as intervention period with practice/training of at least 3x/week was chosen based on prior literature.^5^

Only the first author (principal investigator, VDW), instructors that provided the introduction class, and the participants were aware of who was allocated in each group. Students were involved in in-person data assessment (blinded to study allocation). The patient-reported outcome measures were assessed over Zoom. The MRI scanning was done at the Center of Magnetic Resonance Imaging (University of Minnesota). The MRI technician, biostatistician, and brain imaging expert who analyzed the data were blinded to the group allocation.

### 2.5 Interventions

A TIDieR table describes both interventions side-by-side (***Table 1***). More details about each intervention can be found below.

**Table 1.** Description of interventions using the Template for Intervention Description and Replication (TIDieR)^95^.

| **Brief name** | **Qigong (mind-body approach)** | **P.Volve low-intensity exercises** |
| --- | --- | --- |
| **Why** | Rationale: Qigong is found effective for chronic pain, although the evidence for chronic low back pain is scarce. | Rationale: The low-intensity exercises are similar to what is provided by a physical therapist in standard clinical care. |
| **What materials** | Online Video Spring Forest Qigong™ “5 Element Healing Movement”.<br>No other material needed. | Six online Videos, developed by a licensed physical therapist (Dr. Amy Hoover) who collaborates with the P.Volve Fitness Company. Material: Floor mat + P-ball. |
| **What procedures** | Five gentle horizontal and vertical arm and leg movements performed with guided breathing and focus on body awareness and flow of movements. The five gentle movements are preceded and followed by gentle tapping of different body parts. For more information see Van de Winckel et al. (2022)<sup>54</sup> | Functional fitness and restoration via low-impact exercises focused on core and whole-body strengthening and coordination. |
| **Who** | Qigong Grand Master Chunyi Lin, Founder of Spring Forest Qigong™, developed and demonstrates the five Qigong movements in the “5 Element Healing Movement” video. | A licensed Physical Therapist (Dr. Amy Hoover, PT, DPT) developed the six videos for adults with chronic low back pain. These exercises are demonstrated in the video by two P.Volve trainers. |
| **How** | Access to the video on the Spring Forest Qigong webpage with secure login through StudyID and password.<br>https://www.springforestqigong.com/ | Access to the video on the P.Volve webpage with secure login through StudyID and password.<br>https://www.pvolve.com/ |
| **Where** | At their home or any location of their choice with WiFi connection. | At their home or any location of their choice with WiFi connection. |
| **When, how much** | Introduction class by one of the Qigong masters of the Spring Forest Qigong™ center.<br>12 weeks, at least 3x/week, 1 video/session (approximately 41min per video). | Introduction class by a licensed physical therapist and a trainer from the P.Volve Fitness Company.<br>12 weeks, at least 3x/week, 1 video/session (approximately 41min per video). |
| **Tailoring** | All movements can be done standing, sitting or lying down.<br>Dr. Van de Winckel also developed kinesthetic imagery adaptations for participants who may have difficulty performing certain whole-body movements. More details are available in Van de Winckel et al. (2022)<sup>54</sup><br>The first author (Dr. Van de Winckel) is currently a certified practice group leader and Spring Forest Qigong™ level 5 practitioner (out of 5) in the Spring Forest Qigong curriculum). She was available for questions throughout the study and was not blinded to group allocation. | Participants went through the increasingly more challenging videos at their own pace. The first 6 weeks, participants exercised using videos 1-3; the last 6 weeks, participants exercised using videos 4-6. The first author (Dr. Van de Winckel, licensed PT) was familiar with the P.Volve principles and content of the exercises in the videos, and was available for questions throughout the study. She was not blinded to the group allocation. |
| **Modifications** | None | None |
| **Planned + Actual fidelity assessment** | Time and duration of access to the video were automatically logged and were delivered to Dr. Van de Winckel on a weekly basis.<br>Participants also self-reported the number of minutes they practiced per week.<br>Any deviations to the minimum required practice time and reasons (illnesses, healthcare issues,...) were logged on a weekly basis. | Time and duration of access to the video were automatically logged and were delivered to Dr. Van de Winckel on a weekly basis.<br>Participants also self-reported the number of minutes they practiced per week.<br>Any deviations to the minimum required practice time and reasons (illnesses, healthcare issues,...) were logged on a weekly basis. |

#### 2.5.1 Remote Qigong Intervention

The Spring Forest Qigong™’s “Five Element Healing Movements” practice includes five gentle horizontal and vertical arm and leg movements performed with guided breathing and focus on body awareness and flow of the movements.^53^ More information about the “Five Element Healing Movements” can be found in our protocol paper of Spring Forest Qigong™ in adults with spinal cord injury.^54^ The video was developed by Qigong Grand Master Chunyi Lin, MS in holistic healing, who founded the Spring Forest Qigong™ Center in Minnesota in 1995 after decades of study with some of the most renowned Qigong masters in China.

The movements can be done standing, sitting, or lying down and thus Qigong practice is suitable for adults with cLBP. During the Qigong movements, participants focus their awareness on the body (posture and movement), breathing, and mind (meditation).^55^ Qigong practice improves posture, body awareness, physical and psychological health, and well-being.^53,55^

A Qigong Master from the Spring Forest Qigong™ Center taught the introduction class (6h) either at the Spring Forest Qigong™ Center (Minnesota) or on Zoom. Then, participants received a website link to access the “Spring Forest Qigong™ “Five Element Healing Movements” video (41min), https://www.springforestqigong.com/. They logged in with their ID code and password, given at the start of the study, so that no personal information was transmitted. They did not need any equipment. They were asked to practice individually at home with the “Spring Forest Qigong™ “Five Element Healing Movements” video for 12 weeks, at least 3x/week. The Qigong website automatically monitored the days, time, and duration that videos were accessed, which allowed us to objectively track adherence. The website administrators of the Spring Forest Qigong™ Center provided weekly logs of all participants to the first author (VDW).

The first author (VDW) was at the time of data acquisition Level 3 in the Spring Forest Qigong™ curriculum and she had experience with the Spring Forest Qigong™ “Five Element Healing Movements” practice. She was available for additional calls or Zoom meetings at the request of the participants during the 12 weeks of Qigong practice to address questions about Qigong and demonstrate movements if needed.

#### 2.5.2 Remote low-intensity exercises

The proposed exercise program (P.Volve) encompassed whole-body low-intensity exercises focused on functional restoration, core strengthening and stabilization/coordination exercises. Moreover, the program delivery was similar to Qigong (i.e., through online videos of similar length). The exercises were developed by a physical therapist of the P.Volve team with clinical experience in treating adults with cLBP and extended knowledge in P.Volve exercise principles.

An introduction class was given by a trainer and a physical therapist of the PVolve Team. The class was held in the first author’s Brain Body Mind Lab, at the University of Minnesota. During the introduction class, a sample of the exercises were demonstrated and hands-on guidance was given to ensure proper form during exercises. Online videos for individual home practice were provided on a secure website hosted by P.Volve, https://www.pvolve.com/. Participants logged in with their ID code and password to exercise 41min/session, at least 3x/week for 12 weeks. Participants received a P.ball (i.e., a small ball with elastic bands attached on each side so the legs can be pulled through and the ball is secured right under the pelvis between the legs) and a floor mat. Other exercises were done without equipment. The videos were tailored to adults with cLBP. The six videos were designed with increasing intensity so that the participants could progress through the videos at their own pace over the 12 weeks.

The first author (VDW), who is a physical therapist, was familiar with the P.Volve principles and with all the exercises in the videos proposed in this study. She was available for additional calls or Zoom meetings at the request of the participants during the 12 weeks of the P.Volve low-intensity exercise training to address questions about the exercises and demonstrate movements over Zoom if needed.

### 2.6 Outcomes

MRI scanning and behavioral assessments were acquired at baseline and after 12 weeks of Qigong practice or P.Volve exercise for feasibility purposes. Zoom sessions were held to complete the patient-rated outcome measures. Graduate students performed the in-person assessment of objectively quantifying ankle proprioception. Data were entered into the University of Minnesota’s Research Electronic Data Capture (REDCap) database. REDCap uses a MySQL database via a secure web interface with data checks to ensure data quality during data entry.

#### 2.6.1 Primary outcome measure: Brain imaging

Participants were scanned for 1.5 hours in a Siemens 3-T Prisma scanner at the Center for Magnetic Resonance Research (CMRR) at the University of Minnesota. Structural MRI imaging with T1-weighted magnetization-prepared rapid acquisition with gradient echo (MPRAGE) [repetition time (TR)=2.5s; echo time (TE)=4.5ms; 0.8mm isotropic voxels] and T_2_ weighted sampling perfection with application-optimized contrasts using different flip angle evolution (SPACE) [TR=3.2s; TE=565ms; 0.8mm isotropic voxels] were acquired on each participant.

Resting-state and task fMRI scans were obtained with a T2^*^-weighted multiband echo planar acquisition tipped 30 degrees relative to the anterior commissure–posterior commissure (AC-PC) plane as determined by the Siemens auto-align head software. This acquisition protocol was designed to measure whole-brain blood-oxygen-level-dependent (BOLD)–contrast with optimal temporal and spatial resolution and to reduce signal dropout [TR=0.8 s; TE=37 ms; flip angle=55 degrees; 72 slices; multiband factor 8; 2 mm isotropic resolution].

For the resting-state fMRI imagery (12min 10sec), participants maintained eye fixation with a restful mind and were asked to stay awake.^56^ We selected the parietal operculum (parts OP1/OP4) and insula as region-of-interest (ROI) based on their importance in sensorimotor function, pain, and body awareness.^33,40,41,45,57–63^

The first fMRI task was pain imagery (8min 15sec). Participants focused on their lower back and performed a gentle focused mental body scan to register any sensation of pain. An auditory cue prompted them when to focus on the sensations in the lower back and when to rest.

The second fMRI task was kinesthetic imagery of a whole-body Qigong movement (19min 51sec). The participants first watched a 24sec video demonstration of Grand Master Lin’s standing, and moving his hands away and closer to the body symmetrically with guided breathing while at the same time alternating bending and extending the knees (i.e., the second movement of the Spring Forest Qigong™’s “Five Element Healing Movement”). After the video demonstration, while lying in the scanner and not moving, participants imagined the feeling of performing this movement as if they were upright. This movement was shown and explained to both groups at baseline and repeated at 12 weeks, given that the P.Volve exercise group did not practice this movement. An auditory cue prompted them when to imagine the movements and when to rest.

The third fMRI task was a shape discrimination task with our custom-build fMRI compatible robot (10min 13sec). The protocol of this task has been described in our previous publications.^40,59,61^ The finger is passively moved by the robot.^40,41,59,61^ The design was set up to identify the cognitive process of discriminating shapes (i.e., awareness of finger position and movement). Given that our prior studies^40,41,59,61^ demonstrated that this task activated brain areas relevant for body awareness and pain (i.e., parietal operculum, insula, posterior parietal cortex), we wanted to investigate the feasibility of brain activation changes with Spring Forest Qigong™ or after P.Volve exercises in adults with cLBP.

#### 2.6.2 Primary outcome measure: feasibility indicators and low back pain intensity outcome

Based on models and guidelines for intervention development,^64,65^ and on prior publications related to ***feasibility markers***,^66–69^ we assessed the following *a-priori* feasibility indicators: We estimated feasibility for intervention adherence to be excellent when >80% participants practiced at least 2x/week; and good if ≥70% practiced at least 2x/week. We asked the participants to report weekly on their adherence with the Qigong/P.Volve exercise program (total minutes per week). Both the Qigong and P.Volve online programs recorded minutes of practice automatically on the website, so we could verify the accuracy of the participants’ reporting. Further feasibility benchmarks were: maximum 30% attrition (given that the study started in the midst of the COVID-19 pandemic); none of the questionnaires fully missing in more than 25% of the participants; mild adverse events related to the study of maximum 10% of the participants; and 70% or more participants satisfied with the program. Participants were asked at the end of the study about their satisfaction with the Qigong practice or exercise program.

Given the slow, gentle movements and kinesthetic imagery of Qigong and the low-intensity movements of P.Volve exercises, and weekly check-ins with the participants, the ***risks*** of Qigong practice or P.Volve exercises were considered to be minimal and expected to be limited to mild transient discomfort. A plan was in place to address any (serious) adverse events as per IRB and Good Clinical Practice requirements. If participants were prescribed pain medication prior to the start of the study they were permitted to continue taking those. However, in order to avoid bias from concomitant interventions, other health appointments for low back pain (e.g., osteopathy) were not scheduled or permitted during the duration of the study.

##### As primary behavioral outcome measure

we assessed weekly the highest, average (defined as low back pain level intensity experienced most of the time), and lowest pain intensity levels with the numeric pain rating scale (NPRS).^70^

#### 2.6.3 Secondary outcome measure: proprioceptive acuity

Ankle proprioceptive acuity data were collected with the Ankle Proprioceptive Acuity System (***Figure 1***). The feasibility of this system was established in a previous study.^71^ The testing was performed by graduate students, in person, in the Human Sensorimotor Control Lab (co-author Dr Konczak’s lab).

**Fig. 1.**
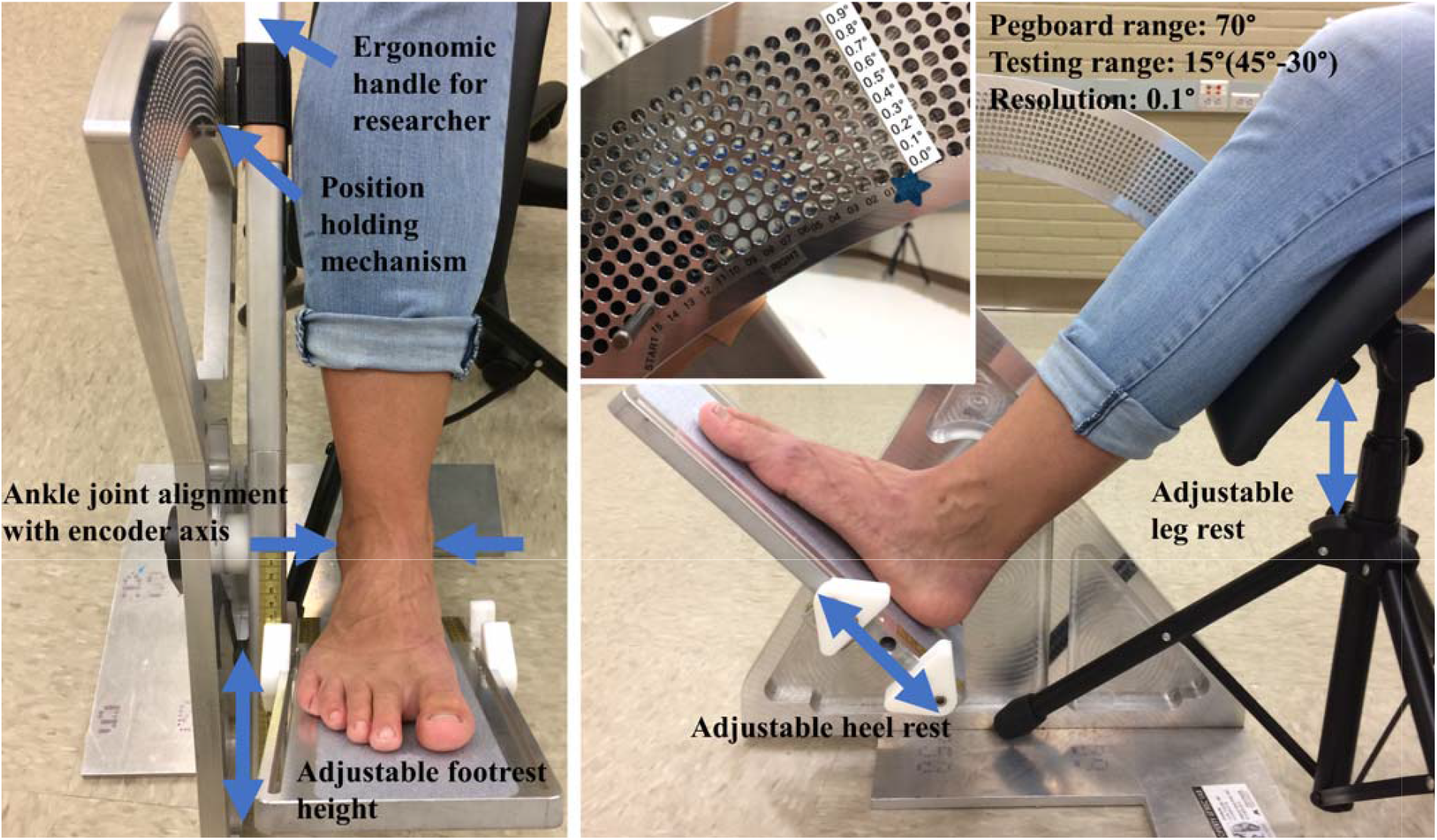
Ankle Proprioceptive Acuity System. To measure ankle proprioceptive acuity in either leg, participants were seated in a chair, barefoot. The participant’s lower leg was supported by an adjustable leg rest, stabilized by a strap at a neutral (90°) joint position. Participants wore vision-occluding goggles.

Participants were seated in a chair, barefoot, to perform the ankle position discrimination in both ankles. Before the testing, the investigator measured the height and length of the lateral malleolus from the heel to adjust the ankle center to align the axis of rotation of the device. The participant’s lower leg was unloaded and supported by an adjustable leg rest, stabilized by a strap at a neutral (90°) joint position (***Figure 1***). Participants wore vision-occluding goggles.

During the testing, the investigator passively plantar flexed the participant’s tested foot from an initial neutral position (90°) to two different plantar flexed positions (i.e., a reference and a comparison position). These two positions were pseudo-randomized during testing. After holding at each position for about 2 seconds, the participant’s foot was then moved back to the initial position. Participants were then asked to verbally indicate in which of the two positions (first or second) their toes were closer to the floor. They were allowed to repeat a trial if needed. Prior to the testing, three practice trials with or without their vision blocked were provided to help participants understand and become familiar with the testing procedure. The practice trials always began with a large position difference (approximately 8°) to help participants understand the goal of the testing. An adaptive psi-marginal algorithm within the predetermined position stimulus range was used so that the next comparison position was based on whether the participant responded correctly to the trial.^72^ A total of 25 trials were performed. Breaks were provided after 10 trials or when the participants requested a break. The total testing took approximately 10-15 minutes to complete.

After 25 trials, a logistic Weibull function was fitted on the stimulus difference (i.e., the difference in reference and comparison ankle positions) and verbal response data for each participant based on the parameter estimates of the psi-marginal adaptive method.^72^ Based on the fitted function, the stimulus difference between the reference and comparison angular positions at the 75% correct response rate was defined as the just-noticeable-difference (JND) threshold, the measure of ankle proprioceptive acuity. A smaller JND threshold (or systematic error) indicates that a person can discriminate smaller angular ankle position differences. The JND threshold represents a marker of ankle position sense acuity.

#### 2.6.4 Secondary outcome measure: patient-reported outcome measures (PROM)

We evaluated body awareness and mindfulness with the following measures: Revised Body Awareness Rating Questionnaire,^73^ Postural Awareness Scale (PAS),^74^ Mindfulness Attention Awareness Scale (MAAS)^75^ and Five-facet mindfulness questionnaire (FFMQ).^76^ MAAS and FFMQ are known to correlate with activation in networks related to mindfulness meditation.^77^

We measured low back disability with the Modified Roland Morris Disability Questionnaire (MRMD),^78^ which measures the degree to which the low back problem or leg pain restricts patients’ daily activities. Function was assessed with the Patient Specific Functional Scale (PSFS).^79^ For this assessment, participants self-identified goals related to activities in daily life that were important to them but that they had difficulty with completing because of the cLBP. The participants rated them between 0 (unable to do the activity) and 10 (able to do the activity).^79^

Core stabilization tests^80^ were done in prone (holding a forearm plank for maximum 2 minutes) and supine bridge positions (holding the bridge position for maximum 2 minutes). We tested balance on each leg^80^ (maximum 2 minutes). The length of time to hold the positions was recorded.

The Fear-Avoidance Beliefs questionnaire^81,82^ assessed high fear avoidance and fear of pain during physical activity (FABQphys-activ) or at work (FABQwork). The Pain Self-Efficacy questionnaire (PESQ)^83–85^ assesses the confidence people have in performing activities despite being in pain. The Tampa Scale for Kinesiophobia (TSK) assesses fear of movement.^86^

Weekly meeting reports of amount of medication used for low back pain, and reports of beneficial effects or adverse events related to the study; and (un)related adverse events (i.e., recent illnesses, health care utilization, and/or recent hospitalizations) were also acquired and entered into REDCap.

### 2.7 Sample size calculation

To our knowledge, this is the first study evaluating the feasibility of brain imaging after a Qigong or exercise intervention in adults with chronic low back pain. This study also provides preliminary estimates of effect size needed to generate hypotheses for future larger randomized controlled trials. We conducted preliminary power calculations using R statistical computing software (*R*).

For the brain imaging outcomes, with n=32 participants, we estimated having 80% power to detect an effect size of at least Cohen’s *d*=1.02 for the difference in brain activation and to detect a change in brain connectivity of at least Δr=0.46, after Qigong or exercise.

For the behavioral outcomes, a decrease in LBP intensity on the VAS have been reported with a within-group effect size of Cohen’s *d*=1.46 when patients practiced Qigong 1x/week for 12 weeks.^30^ Our Qigong practice was more intense (3x/week at home for 12 weeks), so the effect we can detect could be greater.

### 2.8 Statistical analysis

#### 2.8.1 Brain imaging analysis

All neuroimaging data were preprocessed through the Human Connectome Project preprocessing pipeline. We processed the fMRI data using the conn functional connectivity toolbox with established standardized controls for multiple comparisons (SPM family-wise error correction methods).^20^ Data underwent realignment, scrubbing, artifact detection, and CompCor denoising. We focused our primary analyses on the parietal operculum and insula as ROI for resting-state connectivity and exploratory multivariate pattern analyses. Task-based fMRI underwent the same preprocessing pipeline and was modeled with a general linear model.

We used 2 sample t-tests to analyze between-group differences in adults with cLBP compared to 28 healthy adults (scanned in a previous study^52^) of baseline brain function at rest and during tasks. We used paired t-tests for the Qigong group and P.Volve group separately to identify estimates of within-group pre-post changes in task-based brain activation and resting-state connectivity (with Fisher’s Z transformation). We tested the preliminary estimates of effect of Qigong or exercise on brain imaging outcomes with mixed effects models and correlated errors after adjusting for potential confounders, which included the subject-level effect, time, group indicator, and time-by-group interaction as predictors, and other covariates as appropriate. We used Benjamini–Hochberg’s False Discovery Rate (FDR) correction to control for the overall Type 1 errors for the voxel-level analyses.

#### 2.8.2 Clinical assessments

We followed the intention-to-treat protocol. All statistical calculations were performed using *R*. All behavioral primary and secondary outcome measures were analyzed with paired *t*-tests to evaluate pre-post changes in each intervention group (FDR corrected *p*-values and Cohen’s *d* preliminary estimates effect sizes were also calculated to inform future larger randomized controlled trials).

## 3. RESULTS

***Figure 2*** displays the CONSORT study flow chart and reasons for exclusion. In short, of the 36 participants recruited between January 21, 2020 and June 18, 2021, two persons did not pass the screening and two persons withdrew because of conflicting life schedules. The remaining 32 adults with cLBP were enrolled and randomized to either 12 weeks of remote Spring Forest Qigong™ practice or 12 weeks of remote low-intensity P.Volve exercises. Two persons withdrew during the study (Qigong group). In total, 30 adults with SCI completed all study components with the last assessments of this study period completed on September 28, 2021.

**Fig. 2.**
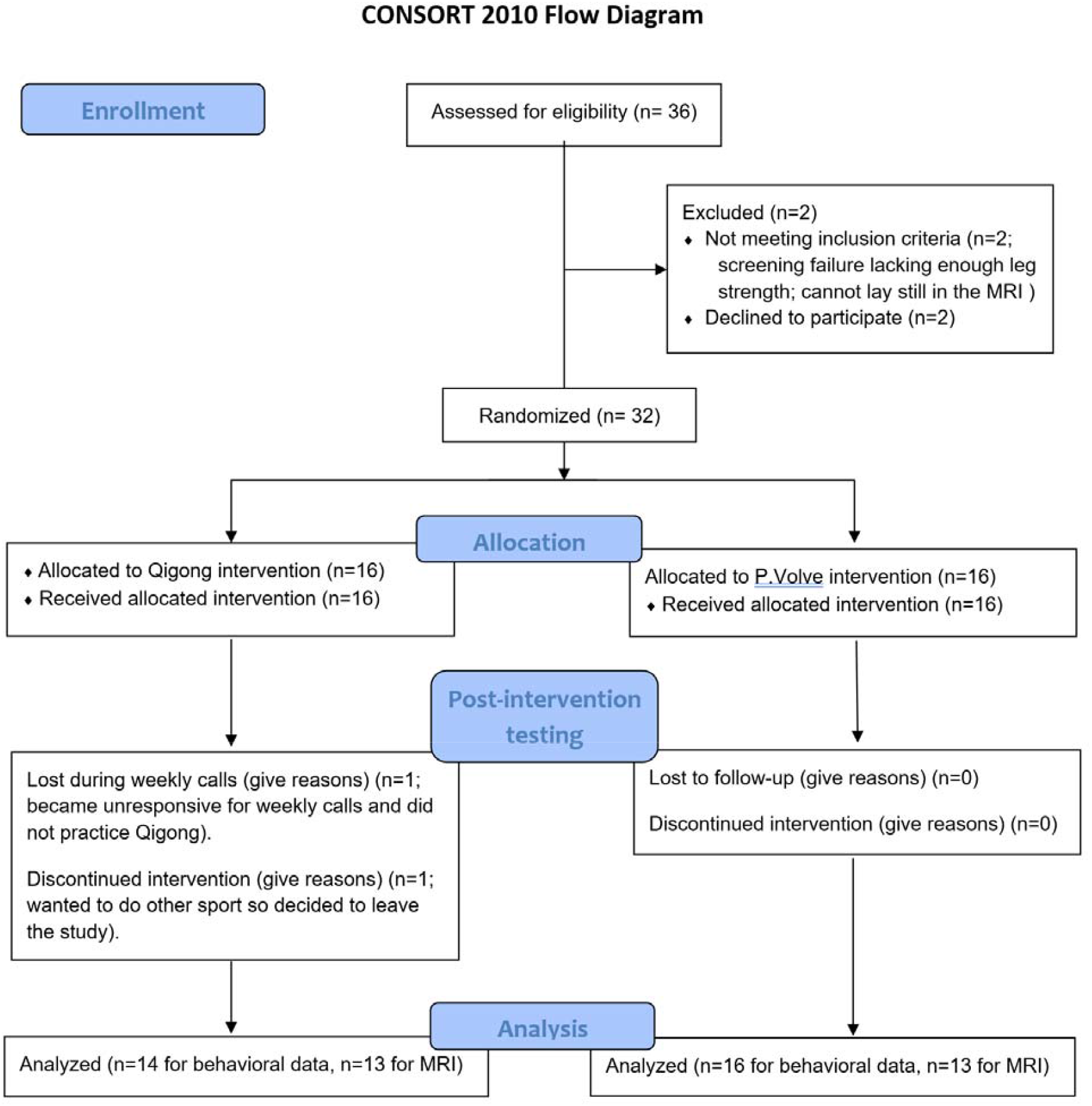
CONSORT flow diagram.

***Table 2*** shows the demographic and clinical characteristics of adults with cLBP per intervention group (n=14 for the Qigong group, n=16 for the P.Volve exercise group), as well as the demographic and clinical characteristics of the healthy adults (n=28), scanned in a previous study.^52^

**Table 2.** Demographic and clinical characteristics of adults with chronic low back pain and of healthy adults (comparison group for MRI testing).

|  | Remote Spring Forest Qigong™ group (n=14) | Remote P.Volve exercise group (n=16) | Healthy adults<sup>52</sup> for MRI comparison (n=28) |
| --- | --- | --- | --- |
| **Age (years)**, mean±SD, range (years) | 46.43±14.87<br>(24-65) | 44.88±15.68<br>(22-70) | 39±16.45<br>(21-69) |
| **BMI**, mean±SD | 25.90±6.28 | 25.52±6.27 | 24.59±3.39 |
| **Sex**, n (%) |  |  |  |
| Male | 2 (14) | 4 (25) | 7 (25) |
| Female | 12 (86) | 12 (75) | 21 (75) |
| Other | 0 (0) | 0 (0) | 0 (0) |
| **Gender Identity** |  |  |  |
| Male | 2 (14) | 4 (25) | 4 (25) |
| Female | 12 (86) | 12 (75) | 11 (39) |
| Other | 0 (0) | 0 (0) | 1 (4) Binary |
| **Ethnicity**, n (%) |  |  |  |
| Hispanic | 1 (7) | 2 (13) | 2 (7) |
| Non-Hispanic | 13 (93) | 14 (88) | 26 (93) |
| Unknown or not reported | 0 (0) | 0 (0) | 0 (0) |
| **Race**, n (%) |  |  |  |
| African American/Black | 0 (0) | 0 (0) | 1 (4) |
| American Indian or Alaska Native | 0 (0) | 1 (6) | 0 (0) |
| Asian | 1 (7) | 1 (6) | 3 (11) |
| Hawaiian or Other Pacific Islander | 0 (0) | 0 (0) | 0 (0) |
| Multi-racial | 0 (0) | 0 (0) | 1 (4) |
| Unknown or not reported | 0 (0) | 0 (0) | 0 (0) |
| White | 13 (93) | 14 (88) | 23 (82) |
| **Baseline chronic low back pain intensity level**, mean±SD |  |  |  |
| High | 6.93±1.33 | 7.50±1.67 | 0.00±0.00 |
| Average | 4.79±1.63 | 4.88±1.75 | 0.00±0.00 |
| Low | 2.14±1.29 | 2.25±1.91 | 0.00±0.00 |
| **Employment Status**, n (%) |  |  |  |
| Disabled due to back pain | 0 (0) | 0 (0) | 0 (0) |
| Disabled for reasons other than back pain | 2 (14) | 0 (0) | 0 (0) |
| Keeping house | 0 (0) | 0 (0) | 0 (0) |
| Looking for work, unemployed | 1 (7) | 0 (0) | 1 (4) |
| Other (volunteer work) | 0 (0) | 1 (6) | 0 (0) |
| Retired | 0 (0) | 2 (13) | 3 (11) |
| Sick leave or maternity leave | 0 (0) | 0 (0) | 0 (0) |
| Student | 1 (7) | 1 (6) | 3 (11) |
| Temporarily laid off | 0 (0) | 0 (0) | 1 (4) |
| Unknown | 0 (0) | 0 (0) | 0 (0) |
| Working now | 10 (71) | 12 (75) | 20 (71) |
| **Education Level, n (%)** |  |  |  |
| No high school diploma | 0 (0) | 0 (0) | 0 (0) |
| High school graduate or GED | 0 (0) | 0 (0) | 0 (0) |
| Some college, no degree | 1 (7) | 2 (13) | 2 (7) |
| Occupational/technical/vocational program | 0 (0) | 1 (6) | 0 (0) |
| Associate degree, academic program | 1 (7) | 2 (13) | 1 (4) |
| Bachelor's Degree | 6 (43) | 6 (38) | 12 (43) |
| Master's Degree | 3 (21) | 4 (25) | 11 (39) |
| Professional school degree | 3 (21) | 1 (6) | 1 (4) |
| Doctoral Degree (PhD) | 0 (0) | 0 (0) | 1 (4) |
| Other | 0 (0) | 0 (0) | 0 (0) |
| **Smoking History, n (%)** |  |  |  |
| Never smoked | 11 (79) | 9 (56) | 25 (89) |
| Current smoker | 1 (7) | 2 (13) | 2 (7) |
| Used to smoke, but now quit | 2 (14) | 5 (31) | 1 (4) |

We obtained excellent intervention adherence as all participants practiced Qigong or did low-intensity P.Volve exercises on average more than 2x/week. More specifically, all of the participants in the Qigong group practiced more than 3 times per week, on average 158.52 minutes per week (128% treatment adherence). The P.Volve exercise group achieved 95.77% treatment adherence. They practiced on average 117.80 minutes per week.

All participants who completed the study enjoyed the practice. There were no study-related adverse events, exceeding our *a priori* benchmark of mild adverse events in max 10% of the participants. All outcomes were collected in all participants at all time points within the accepted time window (within a week of the predefined time points).

We also demonstrated feasibility of the brain imaging resting-state and fMRI tasks. When comparing brain imaging of adults with cLBP with those of healthy adults at baseline, the pain imagery task (***Figure 3 top left picture***) elicited higher activation in the right insula, bilateral parietal operculum, bilateral angular gyrus, and bilateral supramarginal gyrus in healthy adults (n=28) than in adults with cLBP (n=26). Healthy adults had higher precentral gyrus activation than adults with cLBP for the Qigong movement (***Figure 3 top middle picture***) and the robot task (***Figure 3 top right picture***).

**Fig. 3.**
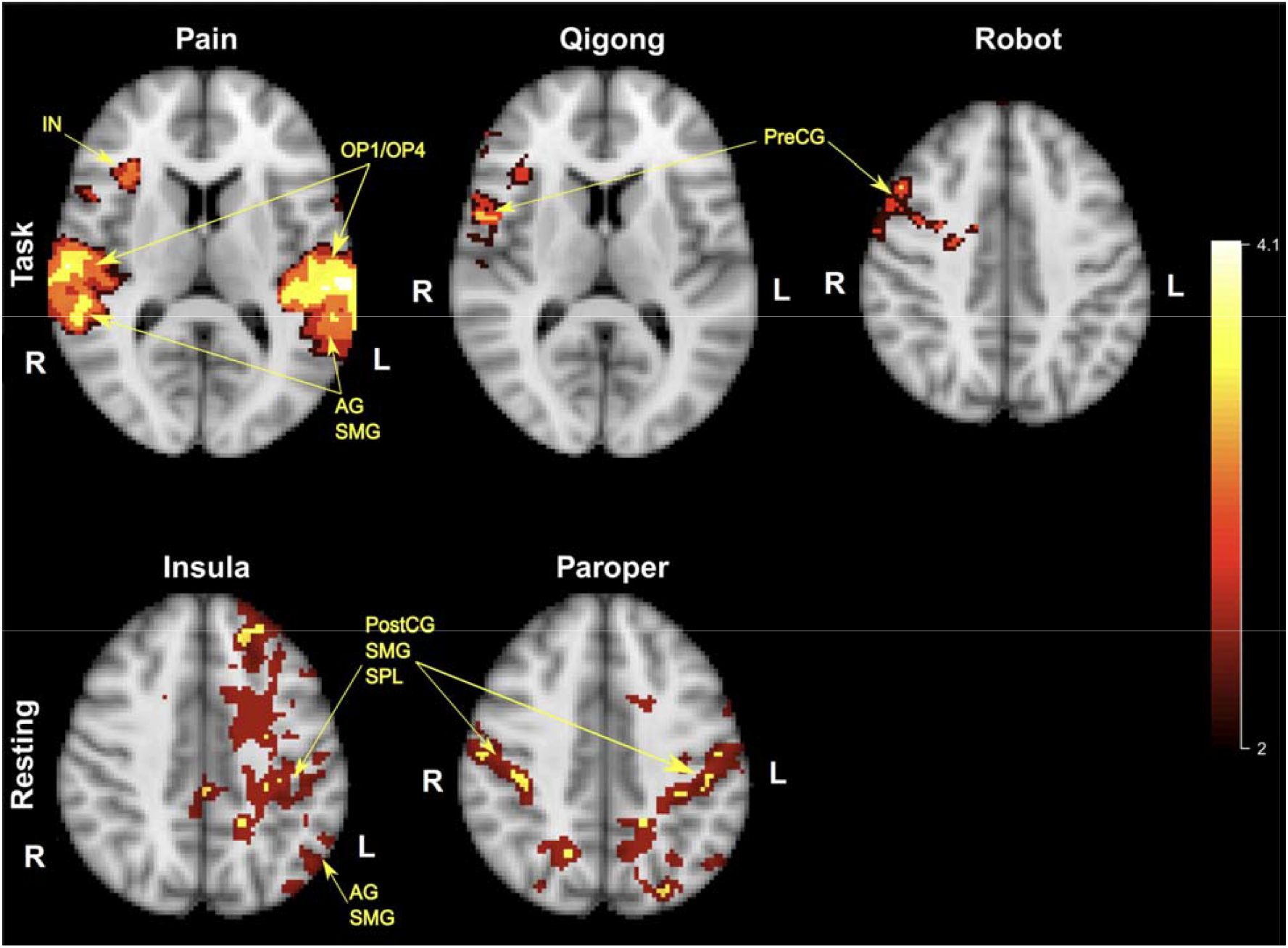
Comparison of brain activation between adults with cLBP (n=26) and healthy controls (n=28) for fMRI tasks (top) and resting-state fMRI (bottom) at baseline. The top left picture shows higher brain activation during the pain imagery task in healthy adults than adults with cLBP (top left picture). Higher activation in the precentral gyrus was seen in healthy adults compared to adults with cLBP for the Qigong task (top middle picture) and for the robot task (top right picture). The bottom left picture shows the insula network connectivity (ROI right insula), and the bottom middle picture shows the parietal operculum network connectivity (ROI right parietal operculum, parts OP1/OP4). We found stronger parietal operculum network and insula network connectivity in healthy adults compared to adults with chronic low back pain. ***Legend:*** AG: angular gyrus; IN: insula; OP1/OP4: parts 1 and 4 of the parietal operculum; PreCG: precentral gyrus; PostCG: postcentral gyrus; SMG: supramarginal gyrus; SPL: superior parietal lobe.

Furthermore, we found stronger parietal operculum network and insula network connectivity in healthy controls (n=28) compared to adults with cLBP (n=26). The ***Figure 3 bottom left picture*** shows stronger connectivity in the insula network between the right insula (ROI) and the left postcentral gyrus, left angular gyrus, left supramarginal gyrus, and left superior parietal lobe. ***Figure 3 bottom middle picture*** shows stronger connectivity in the parietal operculum network between the right parietal operculum, parts OP1/OP4 (ROI) and the postcentral gyrus, supramarginal gyrus, and superior parietal lobe, bilaterally.

Pre-post brain function changes showed individual variability: Six participants in the Qigong group showed increased activation in the parietal operculum, angular gyrus, supramarginal gyrus, and precentral gyrus. Representative examples of 3 participants of the Qigong group are presented in ***Figure 4***, showing brain activation for the Qigong fMRI task at baseline (top row) and brain activation for the Qigong fMRI task after 12 weeks of Qigong practice (bottom row). The bottom pictures show higher activation in the parietal operculum (OP1/OP4), angular gyrus (AG) and supramarginal gyrus (SMG) and additional activation in the precentral gyrus (PreCG) after the 12-week Qigong practice compared to brain activation at baseline.

**Fig. 4.**
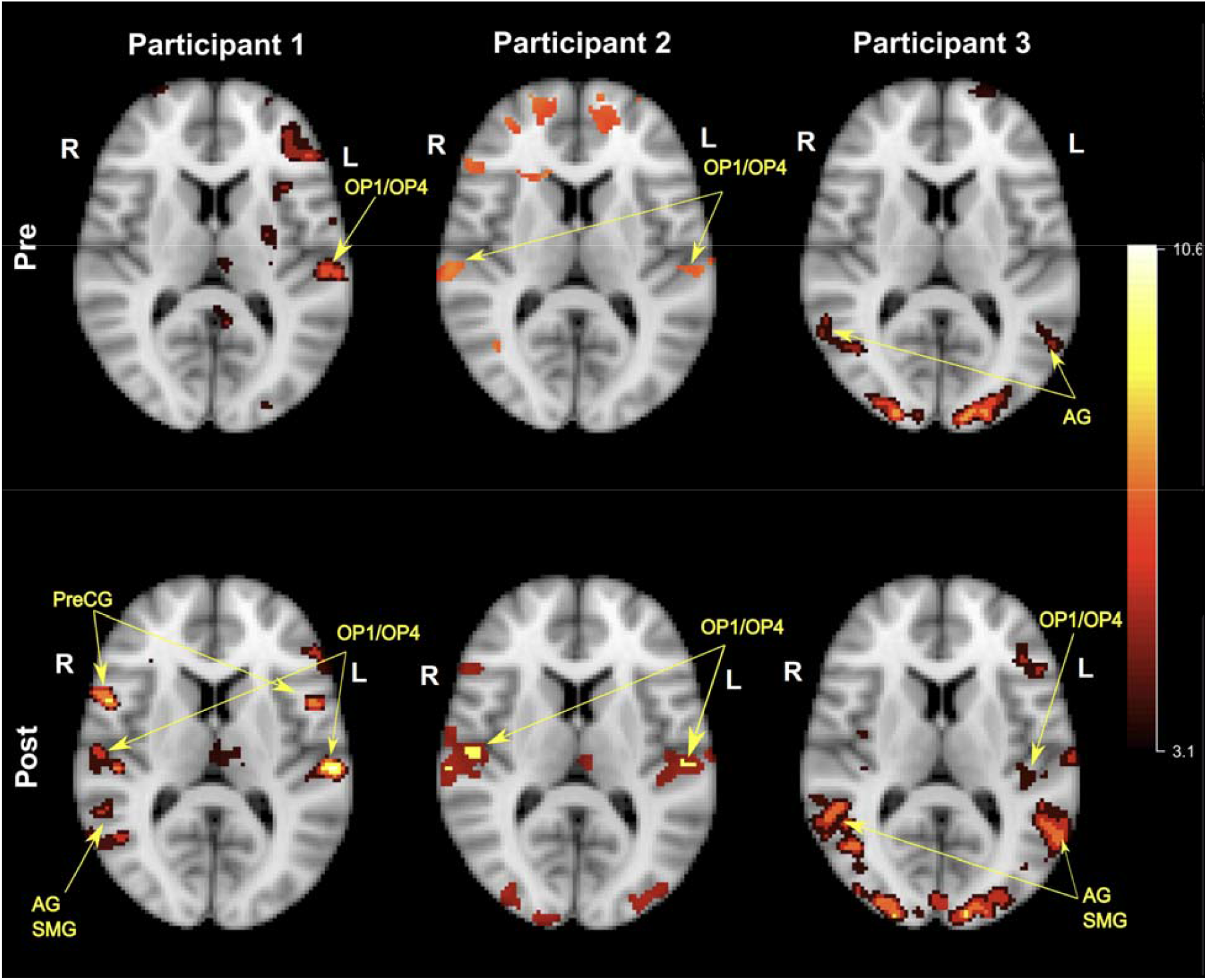
Comparison of brain activation during the Qigong fMRI task between baseline and post-Qigong (12 weeks) in 3 participants with chronic low back pain. *Top figure*: Brain activation during the Qigong fMRI task in three participants at baseline. *Bottom figure*: Brain activation during the Qigong fMRI task in three participants after 12 weeks of Qigong practice. The bottom pictures shows higher activation in the parietal operculum (OP1/OP4), angular gyrus (AG) and supramarginal gyrus (SMG) and additional activation in the precentral gyrus (PreCG) after the 12-week Qigong practice compared to brain activation at baseline.

With regard to the behavioral assessments, complete data sets were collected for all 30 participants. ***Table 3*** is showing the pre-post differences for all outcome assessments in either intervention group with *p*-values corrected for multiple comparisons. Both groups significantly reduced their low back pain. In the Qigong group, the highest pain score was reduced by 3.86±2.11 points (large effect size Cohen’s *d*=1.83), average pain by 3.36±1.86 points (*d*=1.81), and lowest pain by 1.29±0.99 points (*d*=1.30). In the P.Volve exercise group, low back pain intensity was reduced by 5.06±2.57 points (*d*=1.97) for highest pain, 3.88±1.75 points (*d*=2.22) for average pain, and 1.88±1.86 points (*d*=1.01) for lowest pain (***Figure 5***).

**Table 3.** Primary and secondary outcome measures pre-post intervention for the Qigong group and P. Volve group in adults with chronic low back pain.

|  | Remote Spring Forest Qigong<sup>IM</sup> group |  |  |  | Remote P.Volve low-intensity exercise group |  |  |  |
| --- | --- | --- | --- | --- | --- | --- | --- | --- |
| CLINICAL ASSESSMENTS | Baseline (mean±SD) | Post-Qigong (mean±SD) | effect size Cohen's *d* | Adjusted *p*-values | Baseline (mean±SD) | Post-P.Volve (mean±SD) | effect size Cohen's *d* | Adjusted *p*-values |
| n | (n=14) | (n=14) |  |  | (n=16) | (n=16) |  |  |
| NPRS_Highest pain | 6.93±1.33 | 3.07±1.98 | 1.83 | 0.0002* | 7.50±1.67 | 2.44±2.16 | 1.97 | <0.0001* |
| NPRS_Average pain | 4.79±1.63 | 1.43±1.45 | 1.81 | 0.0003* | 4.88±1.75 | 1.00±1.21 | 2.22 | <0.0001* |
| NPRS_Lowest pain | 2.14±1.29 | 0.86±1.17 | 1.30 | 0.0043* | 2.25±1.91 | 0.38±0.72 | 1.01 | 0.0150* |
| PSFS | 2.00±1.54 | 7.88±2.21 | 2.33 | <0.0001* | 1.63±1.44 | 7.71±2.20 | 2.15 | <0.0001* |
| MRMD | 8.93±4.71 | 2.29±2.84 | 1.78 | 0.0003 * | 8.13±3.01 | 1.94±1.61 | 1.87 | <0.0001* |
| REVBA | 18.14±5.46 | 12.14±5.13 | 1.37 | 0.0029 * | 17.94±5.96 | 13.06±4.28 | 0.90 | 0.0341* |
| FFMQ | 125.79±16.90 | 156.57±16.83 | 1.51 | 0.0013* | 139.13±25.72 | 147.63±21.75 | 0.75 | 0.0910 |
| MAAS | 3.62±0.76 | 4.40±0.63 | 1.39 | 0.0027 * | 3.88±1.00 | 4.33±0.89 | 0.68 | 0.1423 |
| PAS | 45.29±13.33 | 65.29±13.37 | 1.22 | 0.0064* | 45.69±15.68 | 61.31±12.75 | 1.45 | 0.0006* |
| FABQphys-activ | 14.21±4.98 | 5.71±6.24 | 1.15 | 0.0094 * | 9.88±6.22 | 6.13±5.04 | 0.76 | 0.0910 |
| FABQ work | 14.79±12.89 | 4.93±7.53 | 0.82 | 0.0629 | 8.38±8.25 | 5.75±7.55 | 0.55 | 0.2195 |
| PSEQ | 45.50±9.28 | 54.71±10.02 | 0.86 | 0.0539 | 49.56±7.15 | 56.25±3.79 | 1.03 | 0.0136* |
| TSK | 38.07±5.86 | 28.36±8.33 | 0.95 | 0.0317* | 30.94±5.95 | 27.63±6.96 | 0.65 | 0.1423 |
| Proprioception average | 2.23±0.92 | 1.70±0.43 | 0.56 | 0.1689 | 1.84±0.65 | 1.78±0.63 | 0.08 | 1.0000 |
| Core muscles - plank | 44.64±34.72 | 73.36±36.25 | 1.25 | 0.0056* | 37.56±25.36 | 78.25±37.07 | 1.28 | 0.0020* |
| Core muscles - bridge | 55.93±44.40 | 85.86±42.36 | 1.15 | 0.0094* | 71.94±41.75 | 107.50±29.01 | 0.78 | 0.0843 |
| Balance average | 73.71±46.41 | 94.25±37.26 | 0.82 | 0.0629 | 81.22±42.68 | 95.41±30.74 | 0.66 | 0.1423 |

**Legend:** Balance average: Time in seconds that a person can balance on one leg, score averaged over both legs; FABQ phys act: Fear Avoidance Beliefs Questionnaire related to physical activity; FABQ related to work: Fear-Avoidance Beliefs questionnaire related to work; FFMQ: Five-Facet Mindfulness Questionnaire; L: left; MAAS: Mindful Attention Awareness Scale; MRMD: Modified Roland-Morris Disability questionnaire; n= sample size; NPRS: Numeric Pain Rating Scale; PAS: Postural Awareness scale; PSEQ: Pain Self-Efficacy Questionnaire; Proprioception average: Proprioceptive acuity averaged over both legs; PSFS: Patient Specific Functional scale (average of 3 functional goals); R: right; REVBA: Revised Body Awareness Rating Questionnaire; TSK: Tampa Scale for Kinesiophobia.
\* significant result

**Fig. 5.**
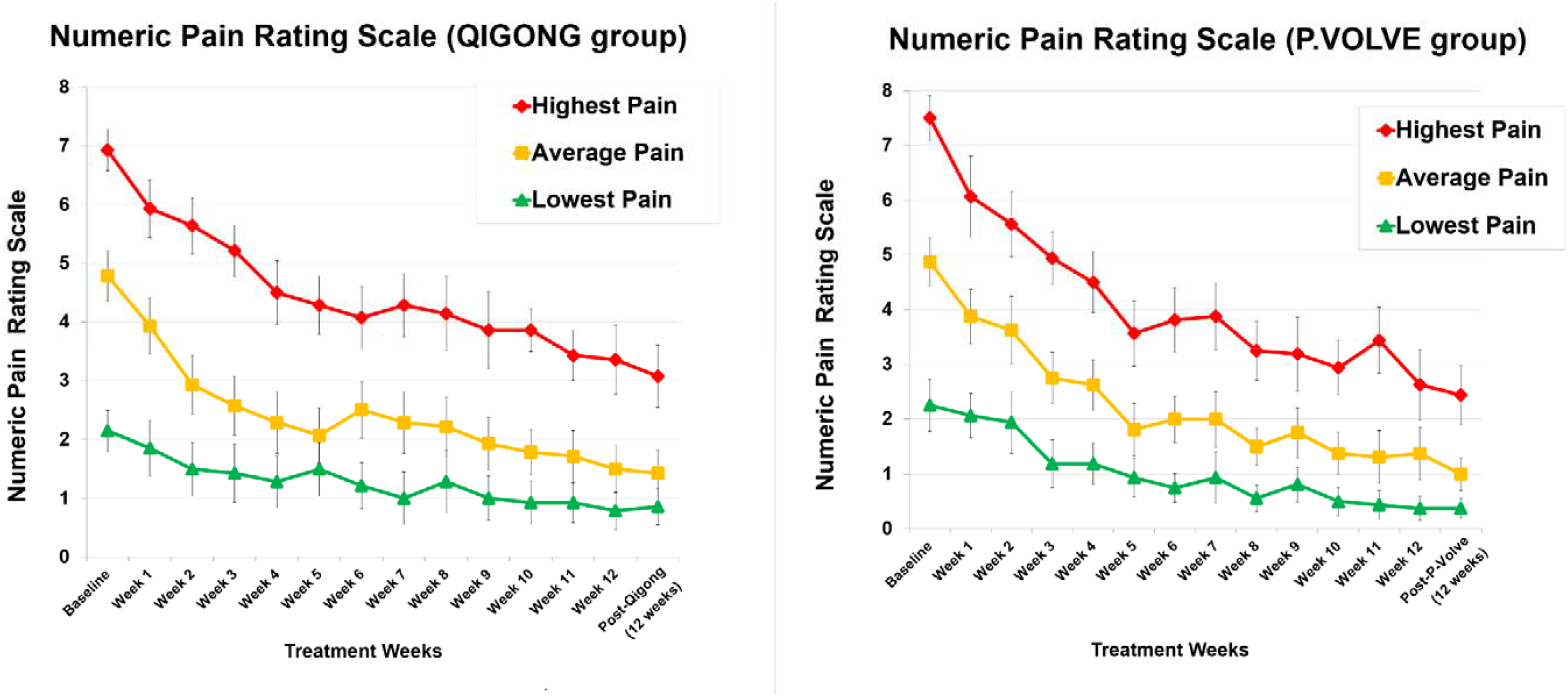
Low back pain intensity ratings in adults with chronic low back pain in the Qigong group and in the P.Volve exercise group. The line graph on the left (Qigong group) and line graph on the right (P.Volve exercises group) represent the weekly highest, average, and lowest low back pain ratings, measured with the numeric pain rating scale.

Significant improvements were also seen for most secondary outcomes with large effect sizes varying from Cohen’s *d*=0.95-2.33 in the Qigong group and *d*=0.90-2.22 in the P.Volve group. Ten outcome measures were significantly improved in the Qigong group. These were measures on disability (MRMD, ***Figure 6***), functional activities (PSFS, ***Figure 7***), body awareness (REVBA, FFMQ, MAAS, PAS), fear of movement (FABQphys-activ, TSK), and core balance measures (holding a plank and a bridge). In the P.Volve exercise group, six outcome measures were significantly improved, related to disability (MRMD, ***Figure 6***), functional activities (PSFS, ***Figure 7***), body awareness (REVBA, PAS), confidence performing activities despite the pain (PSEQ), and core balance measures (holding a plank).

**Fig. 6.**
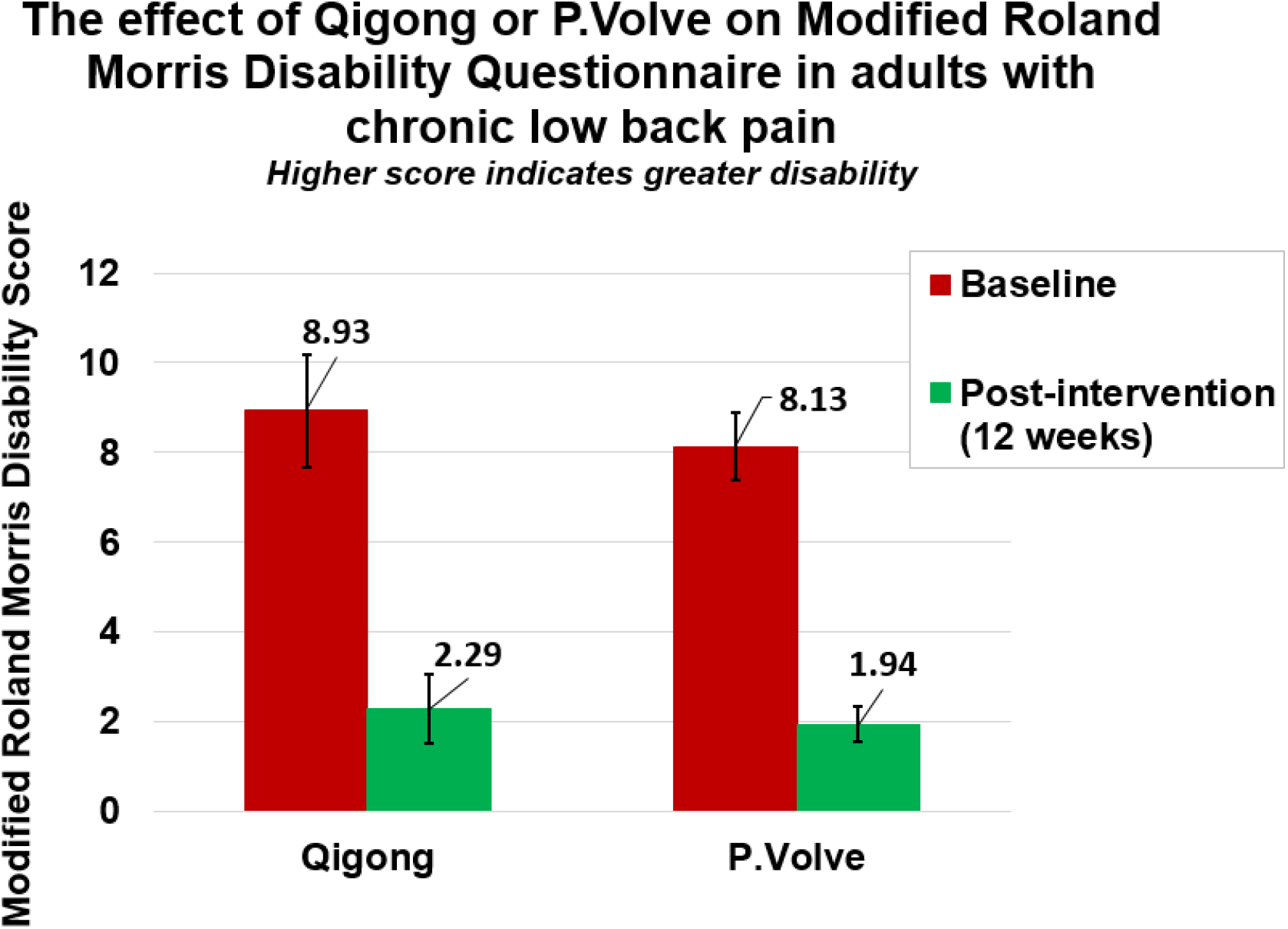
Pre-post intervention improvements on the Modified Roland Morris Disability Questionnaire in adults with cLBP. A higher score indicates greater disability. Both groups show markedly reduced disability after 12 weeks of Qigong or P.Volve exercises.

**Fig. 7.**
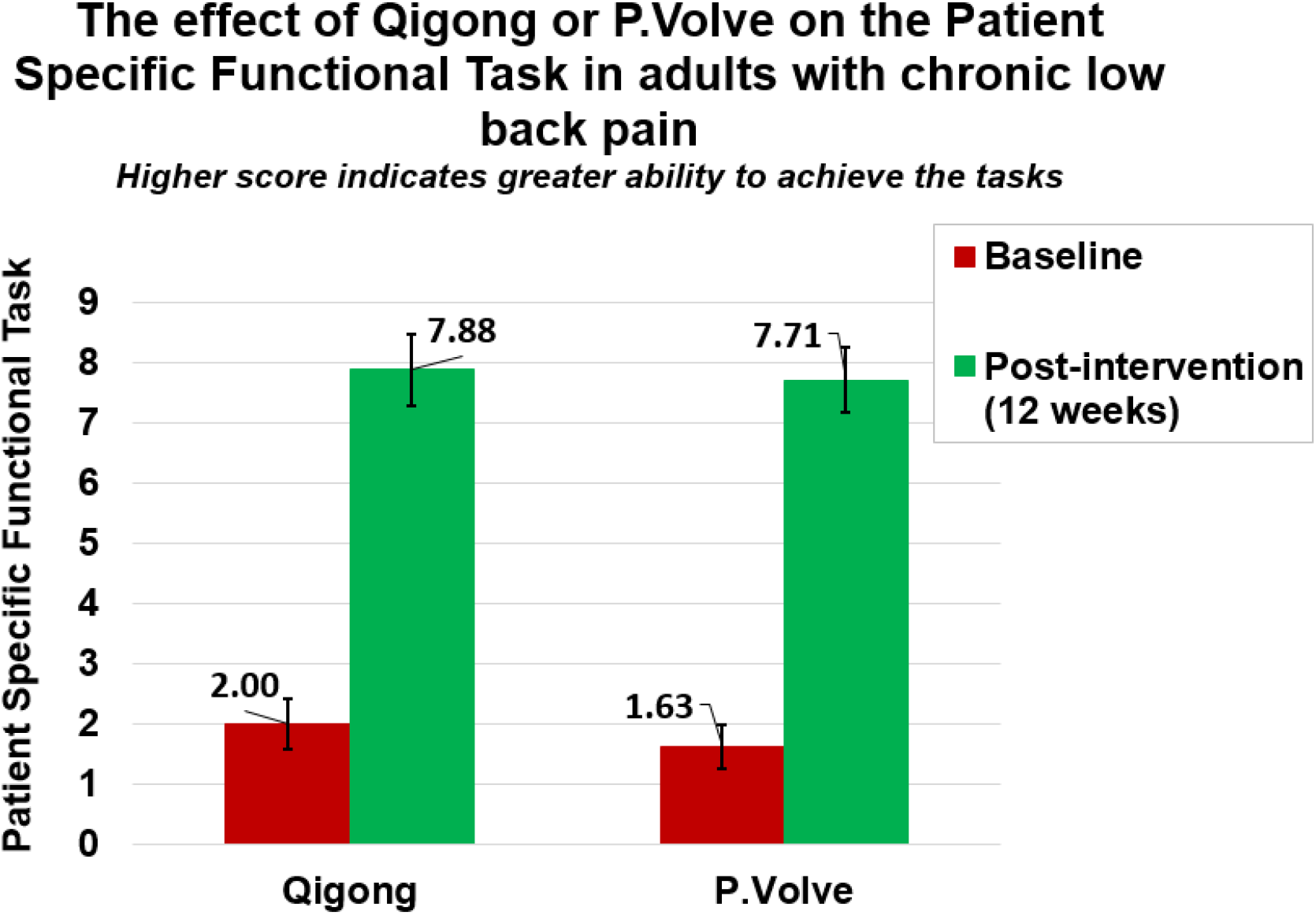
Pre-post intervention improvements on the Patient Specific Functional Task in adults with cLBP. A higher score indicates greater ability to achieve three self-defined functional activities. The bar graphs show an average score of the three activities, rated between 0 (no ability to achieve the functional activities) to 10 (self-defined functional activities are achieved). Both groups improved significantly on their ability to achieve their self-defined functional activities after 12 weeks of Qigong or P.Volve exercises.

## 4. DISCUSSION

The objectives of this study were to investigate the feasibility of practicing remote Spring Forest Qigong™ or remote P.Volve low-intensity exercises, at least 3x/week for 12 weeks, feasibility of recruitment, data collection, delivery of the intervention as intended, as well as identifying estimates of efficacy on brain function and behavioral outcomes after Qigong practice or exercise.^46^ The active control group (P.Volve exercises) were chosen for the similarity to what is provided by a Physical Therapist in standard clinical care and for their focus on functional fitness and restoration low-impact exercises, which were shown to be more effective than other types of exercise treatment for reducing pain intensity and functional limitations.^26^ Both interventions were delivered remotely through online videos of similar duration.

Feasibility clinical trials are important to provide data that are critical for the planning and design of a subsequent clinical efficacy or effectiveness clinical trial. Feasibility markers are the assessment of acceptability of the program, adherence to the dose, frequency, and duration of the interventions, refinement of interventions, selection of appropriate outcomes, ability to capture outcomes within designated timeframes, and/or participant retention.^46,47,87^

We demonstrated the feasibility of our study design and methods, yielding a 94% retention rate. We demonstrated the feasibility of the intervention adherence as adults in the P.Volve group practiced on average between 2-3x/week and adults with cLBP in the Qigong group practiced on average 3-4x/week, thereby exceeding the requested amount of practice of at least 3x/week (128% adherence). We had no missing data in outcomes of those that completed the study (n=30). All participants were engaged and very happy with their respective programs and with the results.

Our results also demonstrated the feasibility of conducting a feasibility randomized controlled trial of remote Spring Forest Qigong™ practice and remote P.Volve exercise training. As estimates of efficacy, both interventions resulted in significant pain reduction and pain-related disability, improved ability to do meaningful functional tasks, improved body awareness, and core strength. The fact that all measures of body awareness and mindfulness were significantly improved in the Qigong group might point to the beneficial combination of present-moment body awareness with gentle movements that the Qigong practice embodies. Participants reported that they felt having greater mental stability, that they were calmer, more centered and resilient, and had a brighter mood. Participants reported that not only their pain was reduced in the lower back but also in other parts of the body. Participants who took pain medication at baseline did not need pain medication after the intervention. At the end of the 12-week Qigong practice, seven adults scored a 1/10 or a 2/10 for highest cLBP intensity in the prior week, and a 1/10 or pain-free most of the time (i.e., average pain intensity), representing 50% of the participants with a very low pain rating at the end of the 12-week Qigong practice. In the P.Volve group, three people were completely pain-free after the intervention, and five either had a 1 or 2/10 for highest pain, and 0/10 or 1/10 for average pain (i.e., pain felt most of the time), also representing 50% of the group with either low pain ratings or no pain after the intervention.

We demonstrated the feasibility of conducting brain imaging (resting-state and fMRI tasks) in adults with cLBP and demonstrated weaker insula and parietal network connectivity in adults with cLBP compared to healthy controls. Lower activation during fMRI tasks in relevant brain areas for body awareness and chronic pain were also seen in adults with cLBP compared to healthy adults. These results extend earlier findings that chronic pain can alter brain function and that insula and parietal operculum areas/networks play a key role in chronic pain perception.^45,88–92^

Interestingly, we saw improved brain activation in six participants after 12 weeks of Qigong in brain areas relevant for sensorimotor function (precentral gyrus), body awareness (parietal operculum) and visuospatial body maps (angular gyrus, supramarginal gyrus). With regard to these results, two elements are worth mentioning: First, further investigation in a larger group is warranted, given that these preliminary findings of improved brain function after Qigong practice are found in 43% of the Qigong group. These preliminary results are in line with studies in other mind and body approaches demonstrating insula activation during meditation practice or changes in the insula brain anatomy and connectivity and higher pain tolerance in long-term yoga practitioners.^93^ Our results are promising because this is the first brain imaging study of Qigong in adults with chronic pain. Second, our findings point to individual cortical signatures of chronic pain processing in adults with cLBP. This observation confirms results found in earlier brain imaging studies in adults with cLBP, pointing to the fact that cLBP is encoded in the anterior insular cortex, the frontal operculum, and the pons; but that, at the individual level, there may be a more complex picture at play. Variability in brain activity is seen across individuals, possibly at least in part explaining the multifaceted aspect of cLBP.^62,94^

The limitations of this study is that we lacked diversity in our sample in terms of sex (the majority of our participants were female), race, and ethnicity. Providing a completely remote study and providing the videos in another language (e.g., Spanish) would help with recruitment of adults from more diverse backgrounds and improve the ability to reach adults with low back pain in rural, underserved areas. Other limitations include the small sample size, which precludes assessment of group differences or assessments of efficacy.

In sum, our pilot data indicate the feasibility and acceptability of using remote Spring Forest Qigong™ practice and remote P.Volve low-intensity exercises for cLBP symptom relief. The remote delivery of Qigong and P.Volve exercises offer multiple applications for broad use in the home or community. The data from the present work will inform the design of future randomized controlled trials with adequate sample size, intervention dosage, and follow-up duration.

This work also demonstrates feasibility of our methods to identify possible mechanisms of how Qigong produces pain relief, and demonstrates individual signatures of chronic musculoskeletal pain processing, pointing to the need for further investigation in larger studies. Having a better understanding of these pain signatures and how Qigong (or exercises) exerts a pain relief effect, could facilitate individually tailored treatments for cLBP.

## Data Availability

All data produced in the present study are available upon reasonable request to the authors.

## Declaration of interests

We declare no competing interests.

## Acknowledgments

This research was supported by the National Institutes of Health’s National Center for Advancing Translational Sciences, grant UL1TR002494, the Biotechnology Research Center: P41EB015894, the National Institute of Neurological Disorders & Stroke Institutional Center Core Grants to Support Neuroscience Research: P30 NS076408; and the High Performance Connectome Upgrade for Human 3T MR Scanner: 1S10OD017974. The content is solely the responsibility of the authors and does not necessarily represent the official views of the National Institutes of Health’s National Center for Advancing Translational Sciences. The funding agency had no role in study design, data collection, data analysis, data interpretation, or writing of the report. Image processing resources were provided by the Minnesota Supercomputing Institute (MSI).

We would like to thank Qigong Master Jaci Gran for teaching the Qigong introduction classes “5 Element Qigong Healing Movements”. Our gratitude goes to Qigong Grand Master Chunyi Lin, his wife Debra Lin for their consultancy and assistance with providing the teaching and content of the “5 Element Qigong Healing Movements”, and to Debra Lin and Collin Silas for the administrative and logical assistance. We would also like to thank Dr. Amy Price Hoover, PT, DPT, and Evan Breed for providing the P.Volve introduction classes and the P.Volve team for the logistical assistance. Finally, we would like to extend our profound thanks to Marc Noël for the critical review of the manuscript.

## Notes

### Competing Interest Statement

The authors have declared no competing interest.

### Clinical Trial

NCT04164225

### Funding Statement

This research was supported by the National Institutes of Health National Center for Advancing Translational Sciences, grant UL1TR002494, the Biotechnology Research Center: P41EB015894, the National Institute of Neurological Disorders & Stroke Institutional Center Core Grants to Support Neuroscience Research: P30 NS076408; and the High Performance Connectome Upgrade for Human 3T MR Scanner: 1S10OD017974. The content is solely the responsibility of the authors and does not necessarily represent the official views of the National Institutes of Health National Center for Advancing Translational Sciences. The funding agency had no role in study design, data collection, data analysis, data interpretation, or writing of the report.

### Author Declarations

The Institutional Review Board (IRB) of the University of Minnesota approved the study (STUDY00005656).

### Summary of Updates

One co-author was inadvertently left out in the first submission.

